# Evaluating the neutralizing ability of a CpG-adjuvanted S-2P subunit vaccine against SARS-CoV-2 Variants of Concern

**DOI:** 10.1101/2021.03.19.21254000

**Authors:** Chia-En Lien, Tsun-Yung Kuo, Yi-Jiun Lin, Wei-Cheng Lian, Meei-Yun Lin, Luke Tzu-Chi Liu, Yu-Chi Chou, Charles Chen

**Affiliations:** Medigen Vaccine Biologics Corporation, Taipei City, Taiwan; Institute of Public Health, National Yang-Ming Chiao Tung University, Taiwan; Department of Biotechnology and Animal Science, National Ilan University, Yilan County, Taiwan; Biomedical Translation Research Center (BioTReC), Academia Sinica; Temple University, Philadelphia, PA 19122, USA

## Abstract

Vaccination is currently the best weapon to control the COVID-19 pandemic. However, an alarming number of novel variants termed Variants of Concern (VoC) were found to harbor mutations that diminished the neutralizing capacity of antibodies elicited by the vaccines. We have investigated the neutralizing titers of antibodies from sera of humans and rats immunized with the MVC-COV1901 vaccine against pseudoviruses coated with the wildtype, D614G, B.1.1.7, or B.1.351 spike proteins. Rats vaccinated with two doses of adjuvanted S-2P retained neutralization activities against the B.1.351 variant, albeit with a slight reduction compared to wildtype. Phase 1 vaccinated subjects showed more reduced neutralization abilities against the B.1.351 variant. The study is among the first, to our knowledge, to demonstrate dose-dependent neutralizing responses against VoCs, particularly against B.1.351, from different doses of antigen in a clinical trial for a subunit protein COVID-19 vaccine. The appearance of vaccine escape variants is a growing concern facing many current COVID-19 vaccines and therapeutics. Strategies should be adopted against the ever-changing nature of these variants. The observations of this study grant us valuable insight into preemptive strikes against current and future variants.

## Introduction

RNA viruses are fast-evolving and can accumulate mutations due to their error-prone polymerases; however, bearing the largest and most complex RNA virus genome, coronaviruses such as the SARS-CoV-2 have proofreading polymerases that allow them to slowly mutate over time to adapt to hosts compared to other RNA viruses^1^. Since the beginning of the COVID-19 pandemic, mutants have been detected periodically. A number of them, termed Variants of Concern (VoC), were found to carry mutations in the crucial receptor-binding domain (RBD) for antibody recognition and neutralization. The most representative of these VoCs, all bearing an N501Y mutation in the spike RBD, are B.1.1.1.7, B.1.351, and P1, which were first reported in the UK, South Africa, and Brazil respectively^2-4^. The implication is the lowered neutralization capabilities of monoclonal antibodies and antibodies induced by vaccines, and these mutations could potentially render the current therapeutics and vaccines ineffective^5^. Major therapeutic and vaccine manufacturers such as Regeneron, Moderna, Pfizer, and AstraZeneca had published reports of variants including B.1.351 and P1 that were highly resistant to neutralization^6-8^. Studies have attributed the resistance to antibody neutralization in B.1.351 and P1 variants to triple mutations K417N, E484K, and N501Y in the spike protein RBD. E484K mutation alone rendering the protein refractory to antibody binding via steric hindrance^6^. The industry has scrambled for strategies to combat these emerging variants, including redesigning the vaccine to elicit either variant-specific or more broadly neutralizing antibodies or administering the additional dose to boost the immune response to compensate for the reduction in neutralization^9-11^.

Medigen’s MVC-COV1901 is a SARS-CoV-2 vaccine consisting of recombinant prefusion stabilized spike protein S-2P with CpG 1018 and aluminum hydroxide (alum) adjuvants which have been shown to induce a very high level of antibody titers in preclinical studies^12^. We sought to ask whether MVC-COV1901 could still defend against these variants with its ability to induce a high level of immunogenicity. We found that antisera taken from the phase 1 vaccinated subjects showed neutralization against the wildtype, D614G, and B.1.1.7 pseudoviruses, while B.1.351 pseudoviruses could be neutralized but to a lesser extent. It was also found that a higher antigen dose elicited better neutralizing antibody response.

## Results

### MVC-COV1901-induced antibodies in rats effectively neutralized variants comparable to the wildtype

We took rat sera from previous toxicology studies to assess antibodies’ neutralization ability against emerging variants and subjected them to wildtype and B.1.351 pseudovirus neutralization assays. As shown in Figure 1, the antibodies were still effective against the B.1.351, although the titers were reduced by 1.49 to 4.6-fold in ID_50_ and 1.4 to 3.05-fold in ID_90_ relative to wildtype. The overall trend is that with the increase of the amount of antigen or adjuvant, the differences between the neutralization titer of wildtype and B.1.351 decrease (Figure 1B).

**Figure 1.**
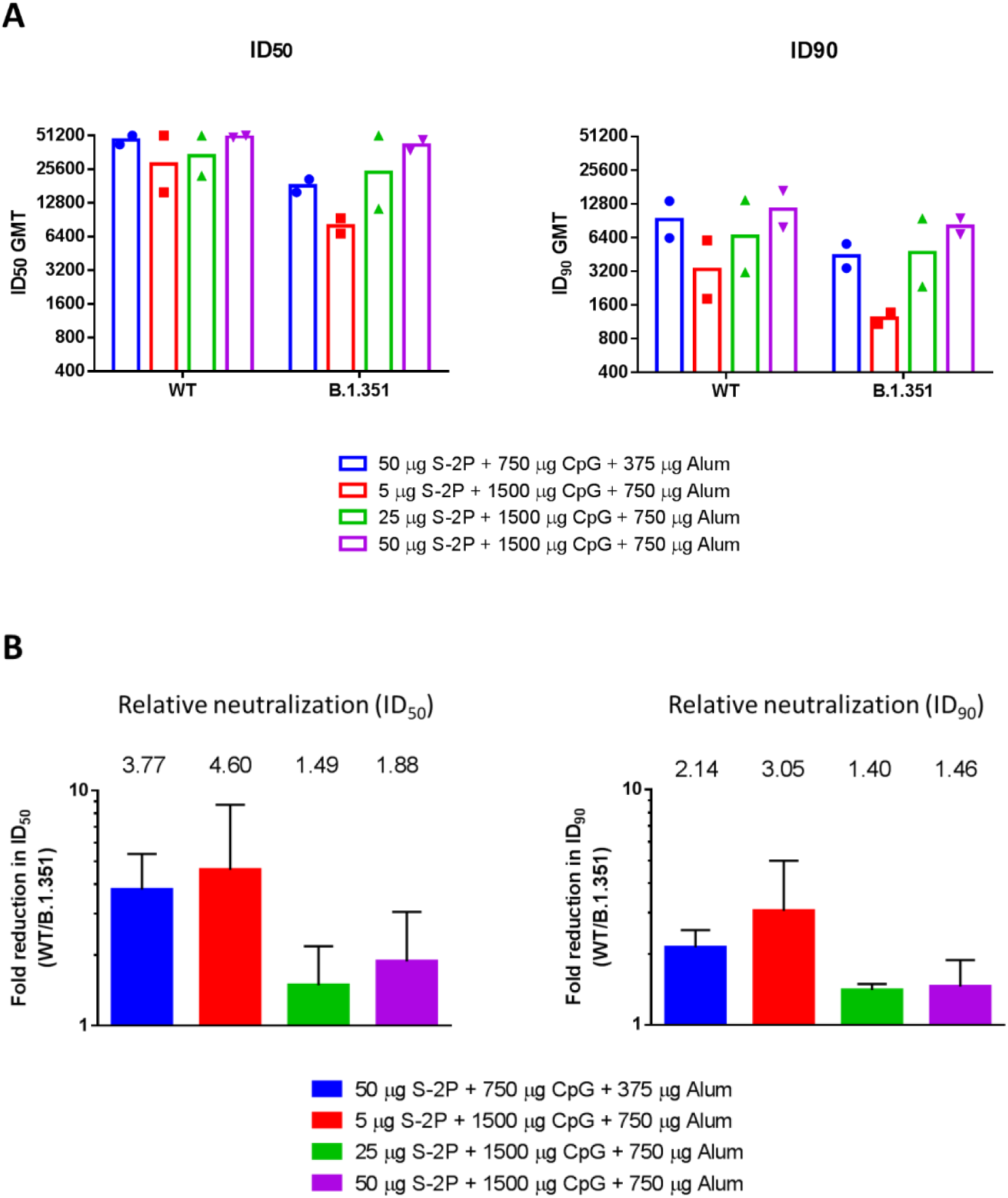
Neutralization of SARS-CoV-2 pseudovirus bearing wildtype or B.1.351 variant spike proteins by antisera of rats vaccinated with adjuvanted S-2P. Rats were immunized twice at 2 weeks apart with the indicated amount of adjuvanted S-2P. For the 50 μg S-2P+750 μg CpG 1018 + 375 μg alum group, 3 males were pooled into one sample and 3 females were pooled into one sample. For the other three groups, 5 males were pooled into one sample and 5 females were pooled into one sample. These resulted in two pooled samples (N=2) for each dose group. (A) The antisera were harvested two weeks after the second injection, pooled as indicated above and subjected to neutralization assay with pseudovirus expressing SARS-CoV-2 Wuhan wildtype or B.1.351 variant spike protein to determine the ID_50_ and ID_90_ titers of neutralization antibodies. (B) Fold reductions in neutralization titers (ID_50_ and ID_90_) were calculated by dividing wildtype neutralization titers by B.1.351 neutralization titers. Results are presented in (A) as geometric mean titers with error bars representing 95% confidence interval and (B) as mean values shown on top of the corresponding bar and error bars representing standard deviation.

In all, we have shown that antibodies elicited by MVC-COV1901 in rats could effectively neutralize the various variants, in particular, the B.1.351 variant.

### Human antisera vaccinated with MVC-COV1901 neutralized D614G and B.1.1.7 variants but is diminished against the B.1.351 variant

After confirming that the antibodies induced by MVC-COV1901 in rats could neutralize the VoCs, we tested human serum samples taken from our phase 1 clinical trial drawn at four weeks after the second immunization with low, mid, or high doses of MVC-COV1901 (S-2P adjuvanted with CpG and alum). Figure 2 presents the data from pseudovirus neutralization assay of human sera with the panel of wildtype, D614G, B.1.1.7, and B.1.351 variants. Although the neutralization titers dropped in D614G and B.1.1.7 compared to the wildtype, the reductions were not statistically significant. However, when comparing B.1.351 with the wildtype, the titers decreased significantly with the median ID_50_ of low, mid, and high dose dropping from 311, 342, and 633 to 36, 53, and 94, respectively (Figure 2A), and ID_90_ of low, mid, and high dose dropped from 80, 85, and 140 to 23, 26, and 36, respectively (Figure 2C). A dose-dependent effect could be observed when plotting each dose group’s neutralizing titers against the variants (Figure 3A). The neutralization titers against B.1.351 could be increased by using a higher dose of antigen. Comparing to the wildtype, B.1.351 resulted in a 10.8 to a 12.8-fold reduction in neutralizing titers for ID_50_ and 4.2 to 5.3-fold reduction in neutralizing titers for ID_90_ (Figure 3B).

**Figure 2.**
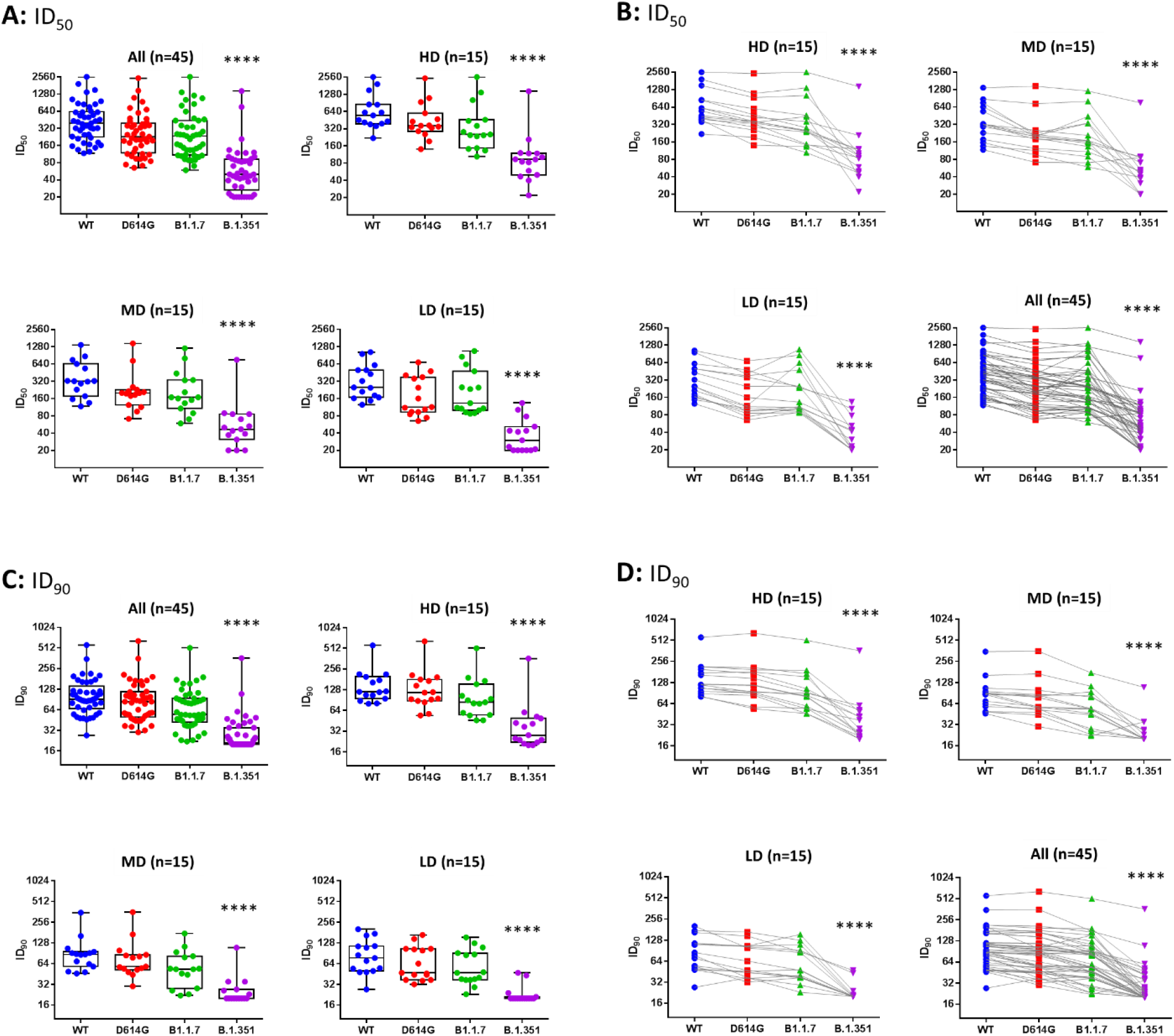
Neutralization of SAR-CoV-2 pseudoviruses with wildtype or variant spike proteins by antisera of clinical trial subjects vaccinated with different doses of MVC-COV1901. Serum samples from clinical phase 1 trial of MVC-COV1901 subjects were collected 4 weeks after the 2^nd^ immunization (56 days from the 1^st^ immunization). (A and B) ID_50_ and (C and D) ID_90_ neutralizing titers for low dose (LD), mid-dose (MD), high dose (HD), and all dose groups were measured with pseudovirus neutralization assays. (A and C) Box plots with median represented by the bar and the box forming the 25% to 75% percentile and whiskers extend to min and max values. (B and D): Results are represented here with each dot representing individual serum sample neutralization titer and lines connect each sample from the WT, D614G, B.1.1.7, and B.1.351 neutralization titers. Kruskal-Wallis, with corrected Dunn’s multiple comparisons test, was used to calculate significance. * = p < 0.05, ** = p < 0.01, *** = p < 0.001, **** = p < 0.0001.

**Figure 3.**
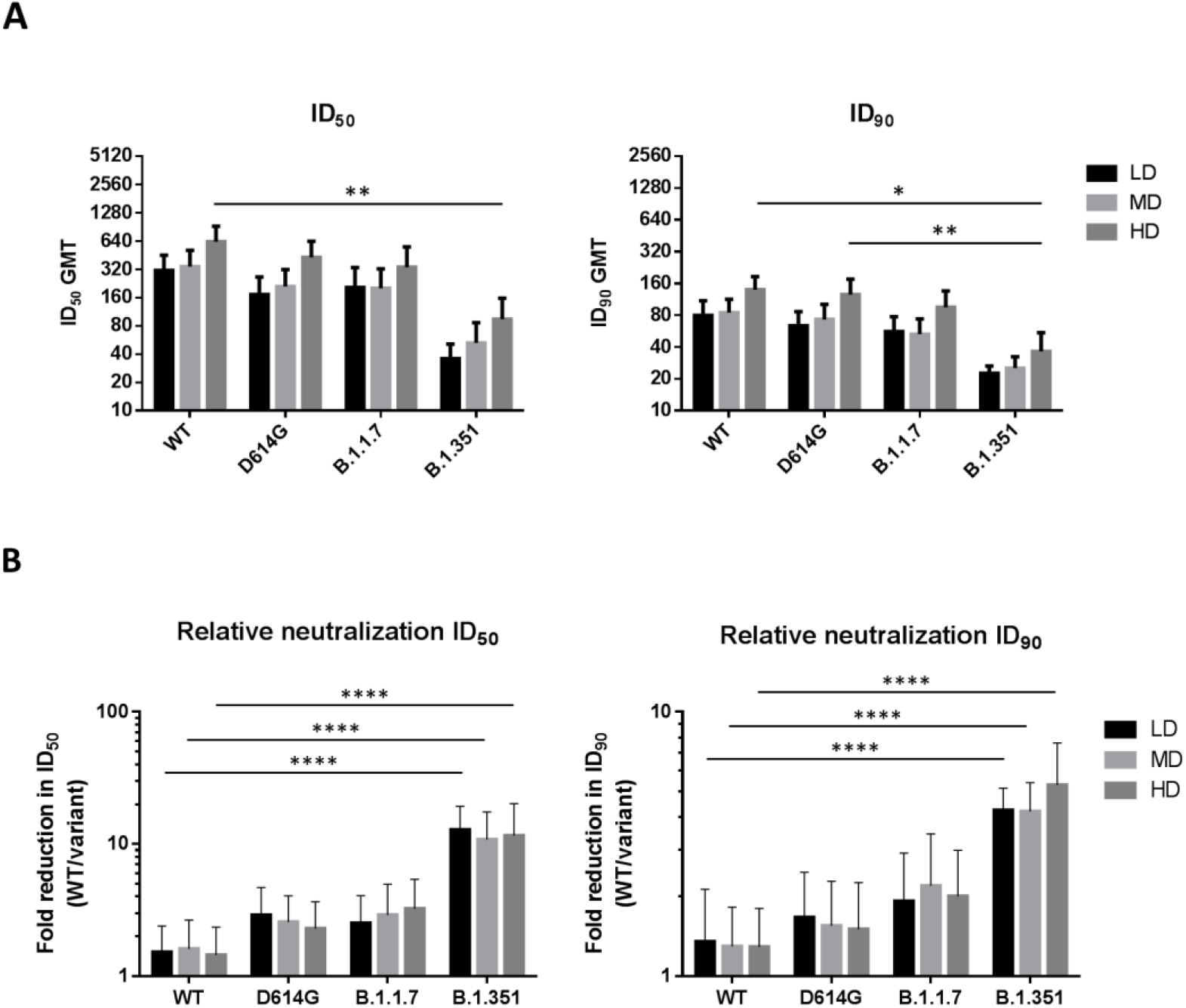
Neutralization of variant pseudoviruses across different doses and fold reduction in the neutralization of D614G, B.1.1.7, and B.1.351 variants relative to the wildtype. (A) ID_50_ and ID_90_ neutralizing titers of each dose group were plotted for the wildtype and variants. The bars and error bars represent geometric mean titers and 95% confidence interval, respectively. (B) Fold reduction in neutralization versus the wildtype was calculated by dividing ID_50_ and ID_90_neutralization titers of wildtype by that of each variant. Results are plotted as bars and error bars indicating mean and standard deviation, respectively. Two-way ANOVA with Tukey’s multiple comparison test was used to calculate significance. * = p < 0.05, ** = p < 0.01, *** = p < 0.001, **** = p < 0.0001.

## Discussion

In this study, we showed that in humans, two injections of a subunit vaccine consisting of the prefusion spike protein (S-2P) adjuvanted with CpG 1018 and aluminum hydroxide were effective in inducing potent neutralization activity against pseudovirus expressing wildtype, D614G and B.1.1.7 variant spike proteins, albeit to a lesser extent to the B1.351 variant. Our study showed that the neutralization titers against B.1.351, compared to the wildtype, reduced by 4.2 to 5.3-fold in neutralizing titers for ID_90,_ comparable to results from studies using Moderna Pfizer’s vaccinee sera as reported by Wang et al. and others^6, 13-14^. The data is also consistent with these studies’ findings, which show slight changes of neutralization titers against the pseudoviruses of D614G or B.1.1.7. We used the sera from participants of our dose-finding, first-in-human clinical trial to obtain neutralizing titers against the pseudoviruses constructed based on the VoCs. With a fixed dose of CpG 1018 and aluminum hydroxide, the data showed that the absolute neutralizing titers against B.1.351 increase with the increase of S-2P antigen in the formulation.

The decreasing difference between the neutralizing titer for wildtype and B.1.351 as the neutralizing tier of wildtype rises is biologically plausible for the below reasons. First, antibody functions other than neutralization have been shown to correlate with protection. The higher the overall neutralizing titer, the larger the reserve of other antibody functions that could be effective against the virus^15-16^. Second, the neutralizing antibodies elicited by MVC-COV1901’s spike protein trimers are polyclonal, as multiple antigenic epitopes could be identified on the protein. The monoclonal antibody (mAb) activities abolished by the mutated site could theoretically be compensated when titers of other neutralizing monoclonal antibodies increase. Amanat et al.^17^ profiled the polyclonal antibodies induced by a SARS-CoV-2 spike mRNA vaccine. They demonstrated the co-dominance of mAbs targeting the N-terminal domain (NTD) and receptor-binding domain (RBD). The mAbs targeting RBD showed smaller abolishment of neutralizing activities against viral variants carrying E484K compared to that of NTD. The competing mAbs bind differentially to variants, suggesting the protective importance of the otherwise-redundant mAbs against the VoCs^18^. It is unclear if the proportion of neutralizing mAbs targeting NTD and RBD would change as the dose escalation was conducted in our first-in-human study, but the overall higher neutralizing antibody titer among the high dose antigen group means a higher dose of effective mAbs against the viral variant in question (Fig. 1 and 2). Our findings suggest that the neutralizing antibody titers elicited by the wildtype strain correlate with the reaction against VoCs. The study results by Garcia-Beltran et al. using sera of 99 vaccinees of Pfizer and Moderna COVID-19 vaccines demonstrated that after the second dose of both vaccines, the neutralizing antibody titers against the pseudoviruses of VoCs increase significantly^5^. The above findings lead to our thinking that it might be a viable option to combat the VoCs by increasing the prototype antigen, the adjuvant, or the number of shots, without redesigning the antigen. Our study generates points of interest warranting further investigation: The possibility of epitope spreading by adjuvant CpG 1018 to extend antibody coverage; the potential of current SARS-CoV-2 vaccine design to protect against the infection of VoCs if the overall neutralizing titer is boosted by the increase of antigen, adjuvant, or the number of shots.

Developing a new COVID-19 vaccine to cope with the VoC is inherently reactive due to the following unknowns, the clinical significance of the emerging strain, the geographical distribution where the strain will be predominant, and how they will mutate further. The FDA has published guidance for the industry to change the antigen based on the VoC to gain regulatory approval using the data accrued from the earlier development stages^19^. Nevertheless, the rollout of variant-specific vaccine that targets a geographical area where the strain is dominant can compromise a vaccination program’s feasibility and timeliness, particularly in a low-and-middle-income setting. Further, it is unknown if the phenomenon of “original antigenic sin” would cause a problem for COVID-19, as a redesigned antigen might preferentially boost the antibodies elicited by the original antigen^20^. Thus, it is desirable to have a COVID-19 vaccine that can cover the emerging variants for a given period before the accumulated mutations lead to a clinically significant abolishment of the neutralizing capacity generated by the vaccine.

Among the emerging VoCs, the B.1.351 variant first reported in the Eastern Cape of South Africa is more problematic. In vitro study has demonstrated immune escapes of the variant from convalescent plasma collected from individuals infected by earlier variants. An attenuation of neutralization titers up to 200 folds with IC_50_ was reported^21^. The implication of these findings at the individual level is possible reinfection by the variant after a previous episode of SARS-CoV-2 infection^22^. It raises the question of whether COVID-19 vaccines that were designed based on the original virus could yield protection against the variant at the public health level. This study investigates how the MVC-COV1901 performs in vitro against the VoCs to evaluate if the MVC-COV1901 can yield clinical protection and inform further development strategy. The study is among the first, to our knowledge, to demonstrate dose-dependent neutralizing responses against VoCs, particularly against B.1.351, from different doses of antigen in a clinical trial for a subunit protein COVID-19 vaccine.

Our study’s limitation is that the sera were taken four weeks after the second shot of MVC-COV19 when the immunity has peaked and starting to wane. Therefore, we could not evaluate the impact of waning immunity on the neutralizing capacity against the variants. The study cannot evaluate the role of T-cell responses elicited by the vaccine as it was reported that the T-cell responses to immunization may confer heterotypic coverage and is less affected by variants of concern^23^. COVID-19 vaccines that conduct phase III clinical trials in South Africa when the B.1.351 strain became predominant showed considerably high protection against severe clinical endpoints, while overall efficacy is lower than sites outside of South Africa^24^. Since there are no correlates of protection being published for either earlier strains or the emerging variants, it warrants further study on how the reduced neutralizing titers will translate to clinical endpoints.

## Methods

### Animal studies

Crl:CD Sprague Dawley (SD) rats were obtained from BioLASCO Taiwan Co. Ltd., and studies were conducted in the Testing Facility for Biological Safety, TFBS Bioscience Inc., and the Center of Toxicology and Preclinical Sciences, QPS Taiwan and immunized as previously described^12^. Briefly, rats were immunized twice at two weeks apart with adjuvanted S-2P with dosing as indicated in Figure 1. The sera were harvested two weeks after the second injection and subjected to neutralization assay with pseudovirus expressing SARS-CoV-2 Wuhan wildtype or B.1.351 variant spike proteins.

All procedures in this study involving animals were conducted to avoid or minimize discomfort, distress, or pain to the animals and were carried out in compliance with the ARRIVE guidelines (https://arriveguidelines.org/). All animal work in the current study was reviewed and approved by the Institutional Animal Care and Use Committee (IACUC) with animal study protocol approval number TFBS2020-010 and CTPS-19-019-01.

### Clinical trial

Forty-five subjects from the age of 20 to 49 were enrolled in a prospective, open-labeled, single-center dose-escalation phase 1 study with three separate sub-phases for participants from 20 to less 50 years of age. Each sub-phase consisted of 15 participants. The three different dose levels employed in this clinical trial are low dose (LD), mid-dose (MD) and high dose (HD) μg of S-2P protein adjuvanted with CpG 1018 and aluminum hydroxide for phase 1a, 1b, and 1c, respectively. The vaccination schedule consisted of two doses, administered by intramuscular (IM) injection of 0.5 mL in the deltoid region of the non-dominant arm, preferably 28 days apart, on Day 1 and Day 29. On Day 57 (4 weeks after the second administration), serum samples were taken for pseudovirus neutralization assays. This phase 1 trial has been registered on clinicaltrials.gov as NCT04487210.

### Pseudovirus neutralization assay

Lentivirus expressing the SARS-CoV-2 spike proteins of the Wuhan-Hu-1 wildtype strain was constructed, and the neutralization assay performed as previously described^12^. Lentiviruses expressing D614G, B.1.1.7, and B.1.351 variant spike proteins were constructed in the same manner but with the spike protein sequence replaced with the respective variant strain.

### Statistical analysis

The analysis package in Prism 6.01 (GraphPad) was used for statistical analysis. Two-way ANOVA with Tukey’s multiple comparison test and Kruskal-Wallis with corrected Dunn’s multiple comparisons test were used to calculate significance as noted in respective figure descriptions. * = p < 0.05, ** = p < 0.01, *** = p < 0.001, **** = p < 0.0001

## Data Availability

The datasets generated during and analyzed during the current study are available from the corresponding author on reasonable request.

## Acknowledgments

We are grateful for the QPS Taiwan and TFBS Biosciences for toxicology studies involving rats. We thank Hao-Yuan Cheng, I-Chen Tai, and Erh-Fang Hsieh of the clinical team for planning and execution of the phase 1 clinical trial.

